# Is Gauchian genotyping of *GBA1* variants reliable?

**DOI:** 10.1101/2023.10.26.23297627

**Authors:** Nahid Tayebi, Jens Lichtenberg, Ellen Hertz, Ellen Sidransky

**Affiliations:** Medical Genetics Branch, National Human Genome Research Institute, National Institutes of Health, Bethesda, MD, United States; Aligning Science Across Parkinson’s (ASAP) Collaborative Research Network, Chevy Chase, MD 20815

## Abstract

Biallelic mutations in *GBA1* result in Gaucher disease (GD), the inherited deficiency of glucocerebrosidase. Variants in *GBA1* are also a common genetic risk factor for Parkinson disease (PD). Currently, some PD centers screen for mutant *GBA1* alleles to stratify patients who may ultimately benefit from *GBA1*-targeted therapeutics. However, accurately detecting variants, especially recombinant alleles resulting from a crossover between *GBA1* and its pseudogene, is challenging, impacting studies of both GD and *GBA1*-associated parkinsonism. Recently, the software tool Gauchian was introduced to identify *GBA1* variants from whole genome sequencing. We evaluated Gauchian in 90 Sanger-sequenced patients with GD and five *GBA1* heterozygotes. While Gauchian genotyped most patients correctly, it missed some rare or *de novo* mutations due to its limited internal database and over-reliance on intergenic structural variants. This resulted in misreported homozygosity, incomplete genotypes, and undetected recombination events, limiting Gauchian’s utility in variant screening and precluding its use in diagnostics.

## Introduction

Gaucher disease (GD) is an autosomal recessively inherited disorder resulting from biallelic mutations in the gene *GBA1* located on chromosome 1q21. *GBA1* encodes the lysosomal enzyme glucocerebrosidase (E.C.3.2.1.45) and more than 500 pathologic variants in *GBA1* have been identified in patients with GD [1]. Variants in *GBA1* are also the most common known genetic risk factor for Parkinson disease (PD) [2-5] The presence of a highly homologous *GBA1* pseudogene, *GBAP1*, located approximately 16kb downstream from the gene, complicates variant detection and sequence analyses [6]. Crossover between two non-sister chromatids during meiosis is an essential cellular process, and highly homologous pseudogenes increase the frequency of nonequal pairing of chromosomes, which can result in complex recombinant alleles [7-9]. While *GBAP1* is 96% homologous to *GBA1* in exonic regions, this sequence similarity increases to ∼98% between intron 8 and the 3’untranslated region (UTR), where a 55bp deletion in exon 9 is the major exonic difference (Figure 1).

**Figure 1.**
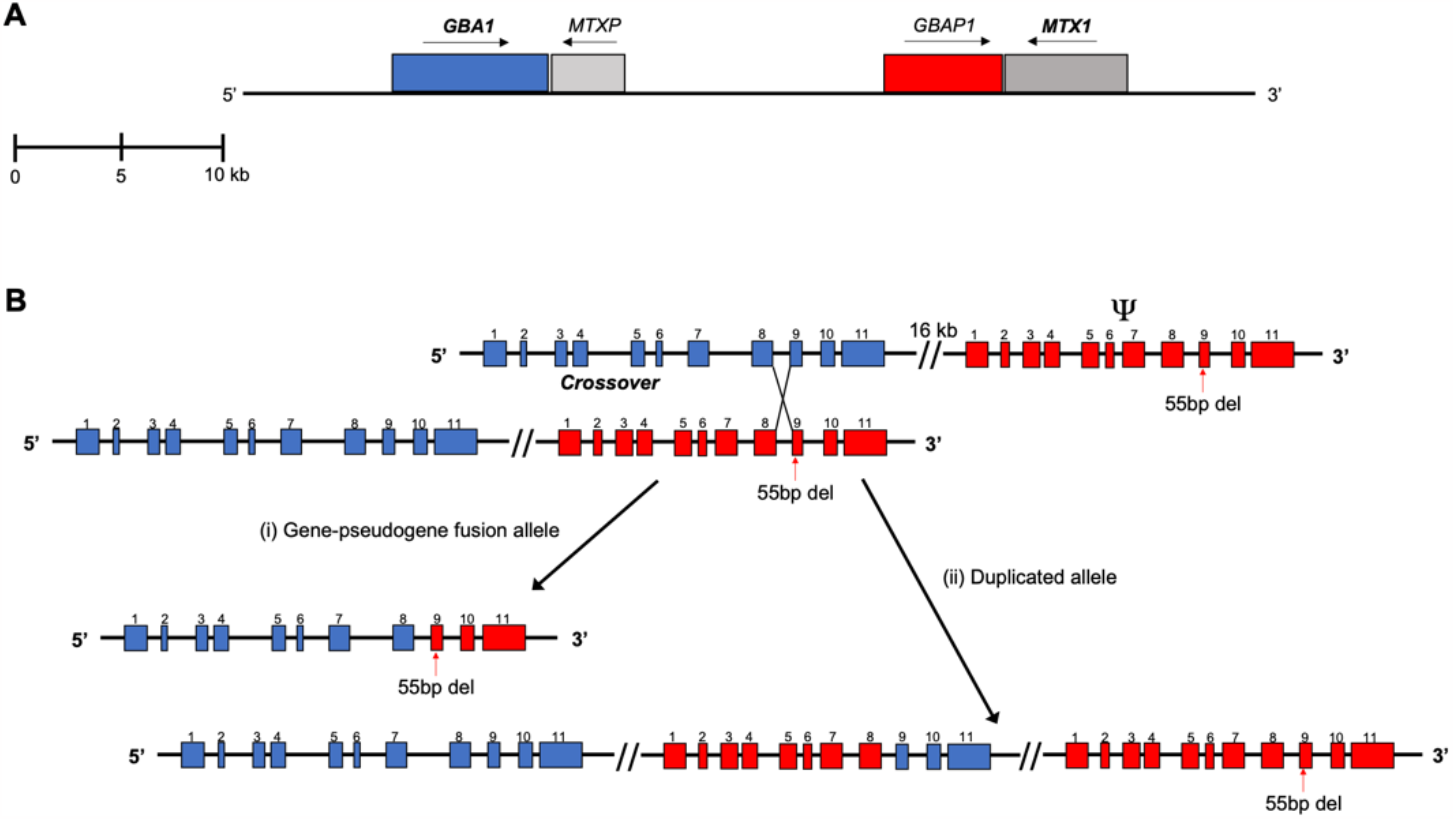
(A) An Illustration of chromosome 1q21 covers two nearby genes and their pseudogenes, *GBA1* and *MTX1*, with sense and antisense. (B) Schematic presentation of a reciprocal cross-over between homologous regions, results in two possible gene rearrangements; one is a fusion between the gene and its pseudogene and a deletion of the intergenic region, the second is a partial duplication of the pseudogene and the gene sequence.

Some Gaucher-related variants are present in *GBAP1* [10, 11]. Furthermore, gene-pseudogene DNA rearrangements comprise a significant proportion of mutant *GBA1* alleles, and such alterations have been detected at different sites between intron 2 to the 3’UTR [12-14]. To date, over twenty different recombinant alleles have been described [15-17], with Rec*NciI* (an allele with three pseudogene SNPs in exon 10), Rec*TL* (Rec*NciI* plus an additional exon 9 pseudogene SNP) and Rec*TL*+55bp (Rec*TL* plus the 55bp del in exon 9) being the most common. While different approaches have attempted to facilitate the computational characterization of the *GBA1* locus [9, 18, 19], consistent identification of recombinant alleles remains a challenge [20]. This impacts both the diagnosis of GD when complex alleles are present, and efforts to quantify and study the impact of *GBA1* in PD [21]. Direct Sanger sequencing, quantitative real-time PCR, Southern blotting and, lately, the correct use of WGS can be used to successfully detect the most common recombinant alleles [12, 15, 22]. Still, these methods can be impractical when analyzing *GBA1* in large populations [23, 24].

Recently Toffoli *et al*. introduced the software tool Gauchian [25] to determine *GBA1* genotypes including point mutations and recombinant alleles from short-read whole genome sequencing (WGS) data. Using sequencing read depth across the 10kb intergenic region between *GBA1* and *GBAP1* as a landmark, Gauchian employs a Gaussian mixture model to call copy number variants, with losses of the intergenic region presumed to represent pathogenic fusion alleles consisting of partial *GBA1* and *GBAP1* fragments, and gains corresponding to an allele with duplication of the 10kb intergenic fragment. The size of the deleted, copy number loss (CNL) or duplicated copy number gain (CNG) fragment depends on the cross-over site between *GBA1* and *GBAP1*, but is not considered further by Gauchian. Gauchian further distinguishes 82 sites that differ between *GBA1* and *GBAP1* to identify the breakpoint between gene and pseudogene variants and uses the 1.1 kb homology region in exons 9 to 11, which contains 10 SNPs and a 55bp deletion in exon 9 of *GBAP1*, to uniquely identify whether a fragment is derived from the gene or pseudogene. This strategy may help characterize *GBA1*/*GBAP1* fusion genes, gene conversions, and duplicated alleles in the region. Due to the ease of use, low performance requirements, and straight-forward result output, Gauchian has already been integrated into many automatic pipelines such as Illumina’s Dragen4, and is incorporated into WGS workflows for neurodegenerative diseases, as it enables quick and easy *in-silico* identification of *GBA1* mutations.

To evaluate the Gauchian tool, we tested a cohort of patients with GD carrying biallelic pathological variants. Both WGS and Sanger sequencing were performed on each DNA sample, and the results were directly compared to the genotypic predictions made by Gauchian.

## Results

We directly compared the genotypic predictions made by Gauchian in 95 samples (190 alleles, although three alleles carried two variants) from our GD cohort, which included patients with and without parkinsonism. Results from WGS were compared to genotypes established via Sanger sequencing (Supplemental Table 1). Gauchian predicted the *GBA1* genotype correctly in 85 samples, while eleven observations did not align with the Sanger/WGS-validated genotypes (Table 1). In three of the eleven cases, Gauchian was unable to establish a fragment copy number count and reported “None”. Genotype visualization generated from WGS and Sanger data for each case misidentified by Gauchian is provided in Supplemental Figure 1 A-J.

**Table 1.**
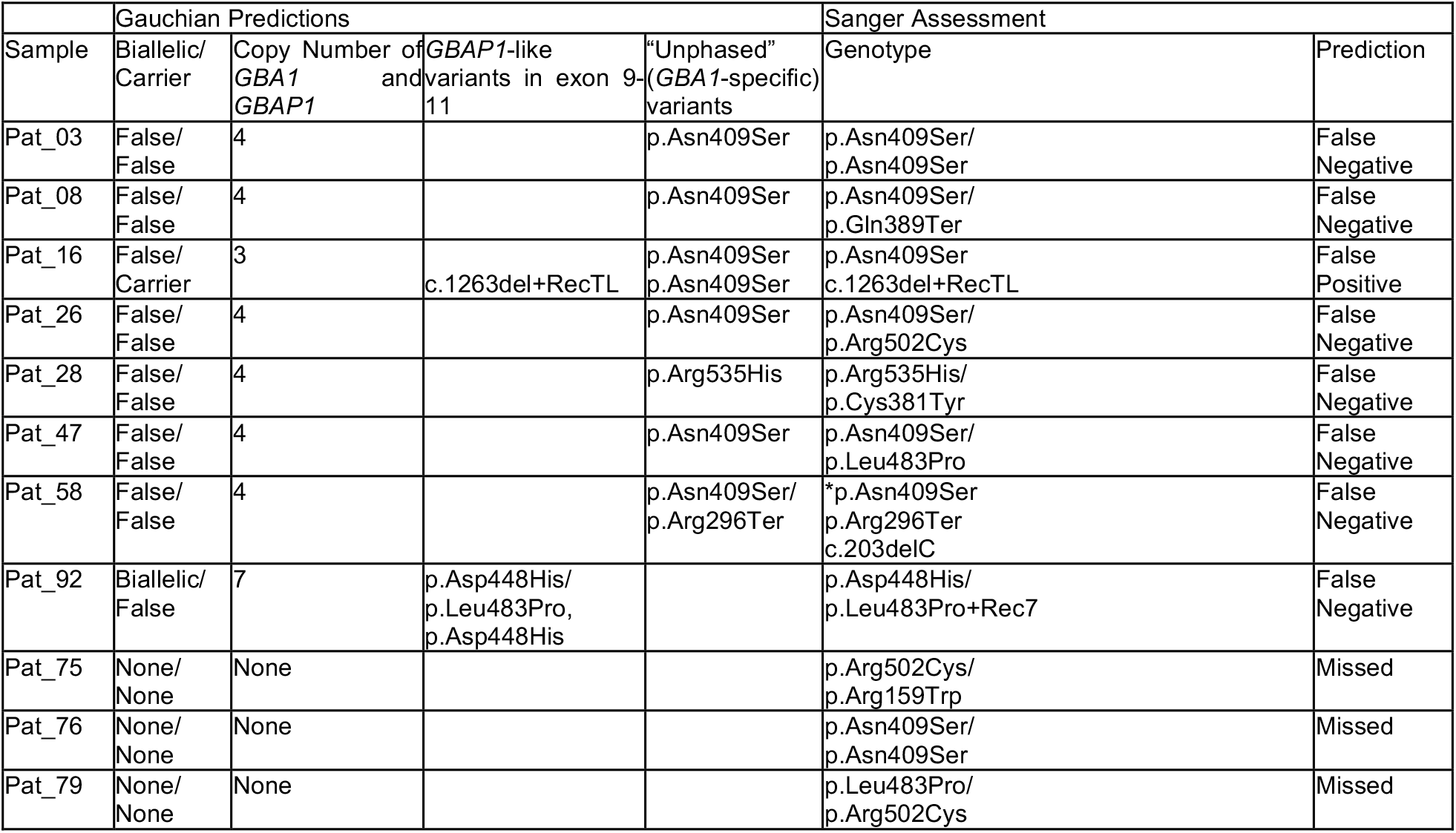
Eleven cases where the Gauchian genotype predictions were not validated by Sanger sequencing. The first four columns represent the output from Gauchian, and the last two columns show the Sanger-established genotype and the prediction assessment for correctness. An asterisk denotes Sanger-established alleles that could not be assigned a phase.

When breaking down the performance of Gauchian based on the individual alleles (Table 2, Supplemental Table 2), of the 193 missense mutations and recombinant alleles characterized by Sanger sequencing, 26 alleles were incorrectly identified, corresponding to an error rate of 13.40%. Of particular concern were eleven alleles erroneously predicted as wildtype alleles. Furthermore, there were instances of failure to detect variants p.Asn409Ser (N370S) and p.Leu483Pro (L444P), which are the most common Gaucher mutations and represent crucial landmarks for clinical genotyping. Also, some rare *GBA1* mutations, e.g., p.Cys381Tyr (C342Y), p.Gln389Ter (Q350X), and p.Gly234Glu (G195E), and some single nucleotide deletions (frameshift mutations) such as c.203del, were not called as they were not annotated in Gauchian’s internal database of known variants. The inability to detect three p.Arg502Cys (R463C) alleles, out of a total of six cases in the cohort, is concerning. The observed failure to correctly identify detect variants and copy numbers in three cases is also problematic, for it would lead to complete misclassification of patients during diagnostic genotyping. Across all alleles characterized by Sanger sequencing, Gauchian achieved a sensitivity of 0.9275 and specificity of 0.9977 (see Supplemental Table 3).

**Table 2.** Gauchian performance regarding individual alleles. True positives (TP) and true negatives (TN), represent the number of correctly identified allele calls (POS) and absences of calls (NEG) for a specific variant, while false positives (FP) represent predictions by Gauchian that have been validated as absent (NEG). False negatives (FN) are omissions that have been validated as observed variants (POS).

| Variant | Benchmark<br>(Sanger) |  | Gauchian<br>(b37) |  |  |  | Gauchian<br>(hg38) |  |  |  |
| --- | --- | --- | --- | --- | --- | --- | --- | --- | --- | --- |
|  | POS | NEG | TP | TN | FP | FN | TP | TN | FP | FN |
| p.Asn409Ser | 122 | 71 | 119 | 70 | 1 | 3 | 114 | 71 | 0 | 8 |
| Rec7 | 1 | 192 | 0 | 192 | 0 | 1 | 0 | 192 | 0 | 1 |
| c.1263del | 3 | 190 | 3 | 190 | 0 | 0 | 3 | 190 | 0 | 0 |
| p.Leu29Alafs*18 | 8 | 185 | 8 | 185 | 0 | 0 | 8 | 185 | 0 | 0 |
| c.203del | 1 | 192 | 0 | 192 | 0 | 1 | 0 | 192 | 0 | 1 |
| p.Cys381Tyr | 1 | 192 | 0 | 192 | 0 | 1 | 0 | 192 | 0 | 1 |
| p.Asp448His | 2 | 191 | 2 | 191 | 0 | 0 | 2 | 191 | 0 | 0 |
| p.Phe252Ile | 1 | 192 | 1 | 192 | 0 | 0 | 1 | 192 | 0 | 0 |
| p.Gly234Glu | 1 | 192 | 1 | 192 | 0 | 0 | 1 | 192 | 0 | 0 |
| p.Gly241Arg | 1 | 192 | 1 | 192 | 0 | 0 | 1 | 192 | 0 | 0 |
| p.Gly416Ser | 1 | 192 | 1 | 192 | 0 | 0 | 1 | 192 | 0 | 0 |
| c.115+1G>A | 3 | 190 | 3 | 190 | 0 | 0 | 3 | 190 | 0 | 0 |
| p.Leu483Pro | 20 | 173 | 17 | 173 | 0 | 3 | 17 | 173 | 0 | 3 |
| p.Gln389Ter | 1 | 192 | 0 | 192 | 0 | 1 | 0 | 192 | 0 | 1 |
| p.Arg159Trp | 2 | 191 | 1 | 191 | 0 | 1 | 1 | 191 | 0 | 1 |
| p.Arg209Cys | 1 | 192 | 1 | 192 | 0 | 0 | 1 | 192 | 0 | 0 |
| p.Arg296Gln | 1 | 192 | 1 | 192 | 0 | 0 | 1 | 192 | 0 | 0 |
| p.Arg296Gln | 1 | 192 | 1 | 192 | 0 | 0 | 1 | 192 | 0 | 0 |
| p.Arg324Cys | 1 | 192 | 1 | 192 | 0 | 0 | 1 | 192 | 0 | 0 |
| p.Arg398Ter | 1 | 192 | 1 | 192 | 0 | 0 | 1 | 192 | 0 | 0 |
| p.Arg502Cys | 6 | 187 | 3 | 187 | 0 | 3 | 4 | 187 | 0 | 2 |
| p.Arg535His | 2 | 191 | 2 | 191 | 0 | 0 | 2 | 191 | 0 | 0 |
| c.1263del+RecTL | 1 | 192 | 1 | 192 | 0 | 0 | 1 | 192 | 0 | 0 |
| RecNcil | 2 | 191 | 2 | 191 | 0 | 0 | 2 | 191 | 0 | 0 |
| p.Val391Leu | 1 | 192 | 1 | 192 | 0 | 0 | 1 | 192 | 0 | 0 |
| p.Val433Leu | 2 | 191 | 2 | 191 | 0 | 0 | 2 | 191 | 0 | 0 |
| p.Arg296Gln | 1 | 192 | 1 | 192 | 0 | 0 | 1 | 192 | 0 | 0 |
| WT | 5 | 188 | 5 | 177 | 11 | 0 | 6 | 171 | 16 | 0 |

**Table 3.** Patients with recombinant alleles and those predicted to have abnormal *GBA1+GBAP1* copy numbers. The first five columns represent the output from Gauchian, and the last column shows the Sanger established genotype.

|  | Gauchian Prediction |  |  |  |  | Sanger Assessment |
| --- | --- | --- | --- | --- | --- | --- |
| Sample | Biallelic/<br>Carrier | Copy Number<br>of<br>*GBA1* and<br>*GBAP1* | *GBA1*<br>Deletion<br>Breakpoint | *GBAP1*-like<br>Variants in<br>Exons 9-11 | "Unphased"<br>(*GBA1*-specific)<br>Variants | Genotype |
| Pat_16 | False/<br>Carrier | 3 | True | c.1263del+RecTL | p.Asn409Ser<br>p.Asn409Ser | p.Asn409Ser<br>c.1263del+RecTL |
| Pat_71 | False/<br>Carrier | 4 |  | RecNcil |  | RecNcil<br>WT |
| Pat_95 | False/<br>Carrier | 4 |  | RecNcil | p.Asn409Ser | p.Asn409Ser<br>RecNcil |
| Pat_42 | False/<br>False | 6 |  |  | p.Arg398Ter<br>p.Val391Leu | p.Arg398Ter<br>p.Val391Leu |
| Pat_72 | False/<br>False | 5 |  |  | p.Gly241Arg | p.Gly241Arg<br>WT |
| Pat_92 | Biallelic/False | 7 |  | p.Asp448His<br>p.Leu483Pro,<br>p.Asp448His |  | p.Asp448His<br>p.Leu483Pro+Rec7 |

Since Gauchian supports the analysis of sequencing data aligned to different references of the human genome and previous work by Pan et al. has shown significant differences between single nucleotide variants using different reference genomes [26], we compared the performance of the software with both b37 (hg19) and hg38 references as a baseline (Supplemental Table 4). While the majority of predictions were congruent, there were four cases with distinct differences. In Pat_16, Sanger sequencing showed a heterozygous p.Asn409Ser mutation. When b37 was used as the reference, Gauchian predicted a homozygous p.Asn409Ser genotype, and with hg38, no mutation was identified. For Pat_35 and Pat_78, Gauchian correctly identified a homozygous p.Asn409Ser genotype using b37 as a reference but missed both mutant alleles entirely when using hg38. For Pat_75, Gauchian missed both Sanger identified variants (p.Arg502Cys, p.Arg159Trp) when using b37 but reported the p.Arg502Cys allele correctly with hg38. There were further discrepancies between the copy numbers reported for Pat_35 and Pat_75 when running the different references. For both patients, hg38 reported CN=3, while b37 predicted CN=4 in Pat_35 and made no call for Pat_75.

**Table 4.**
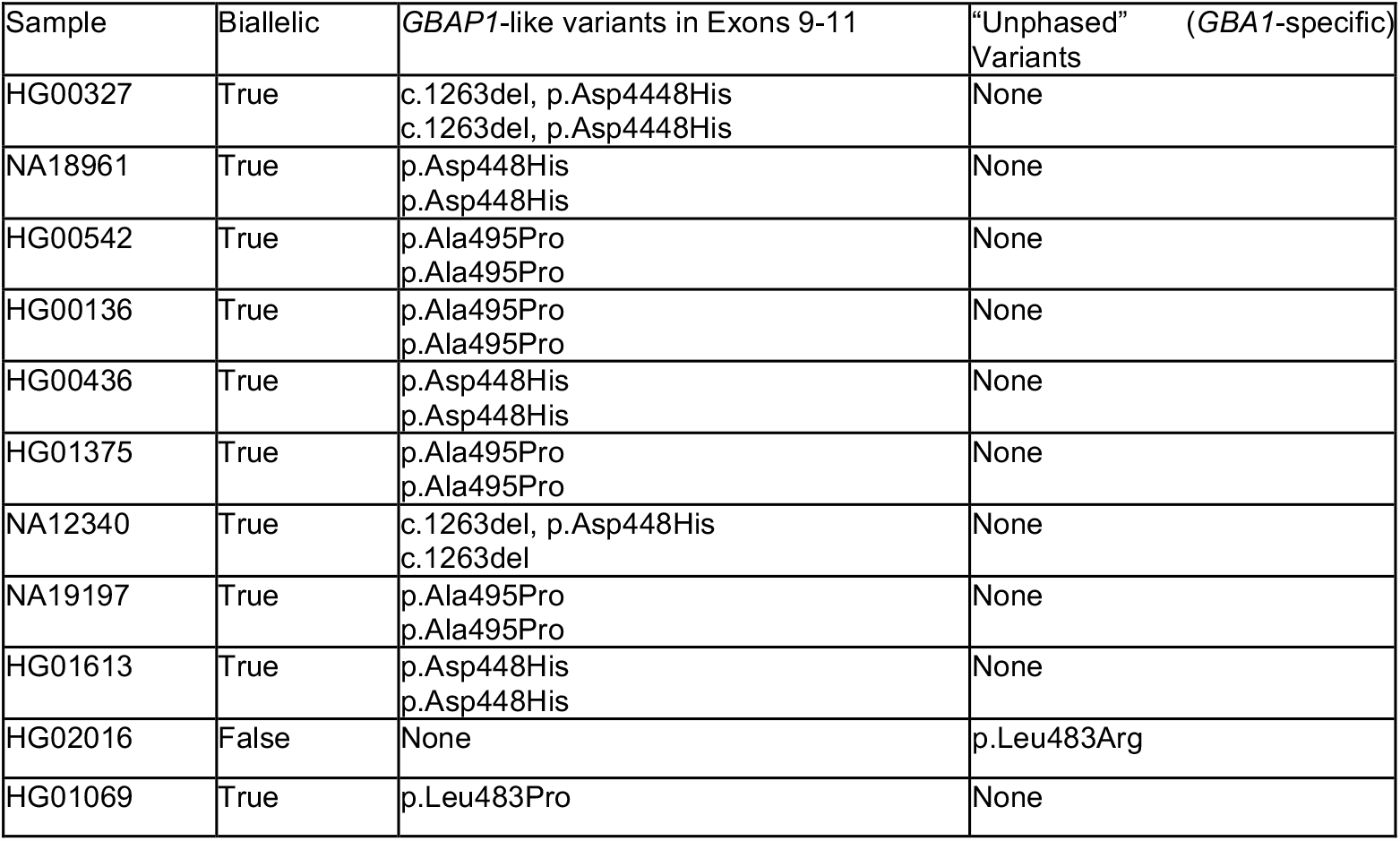
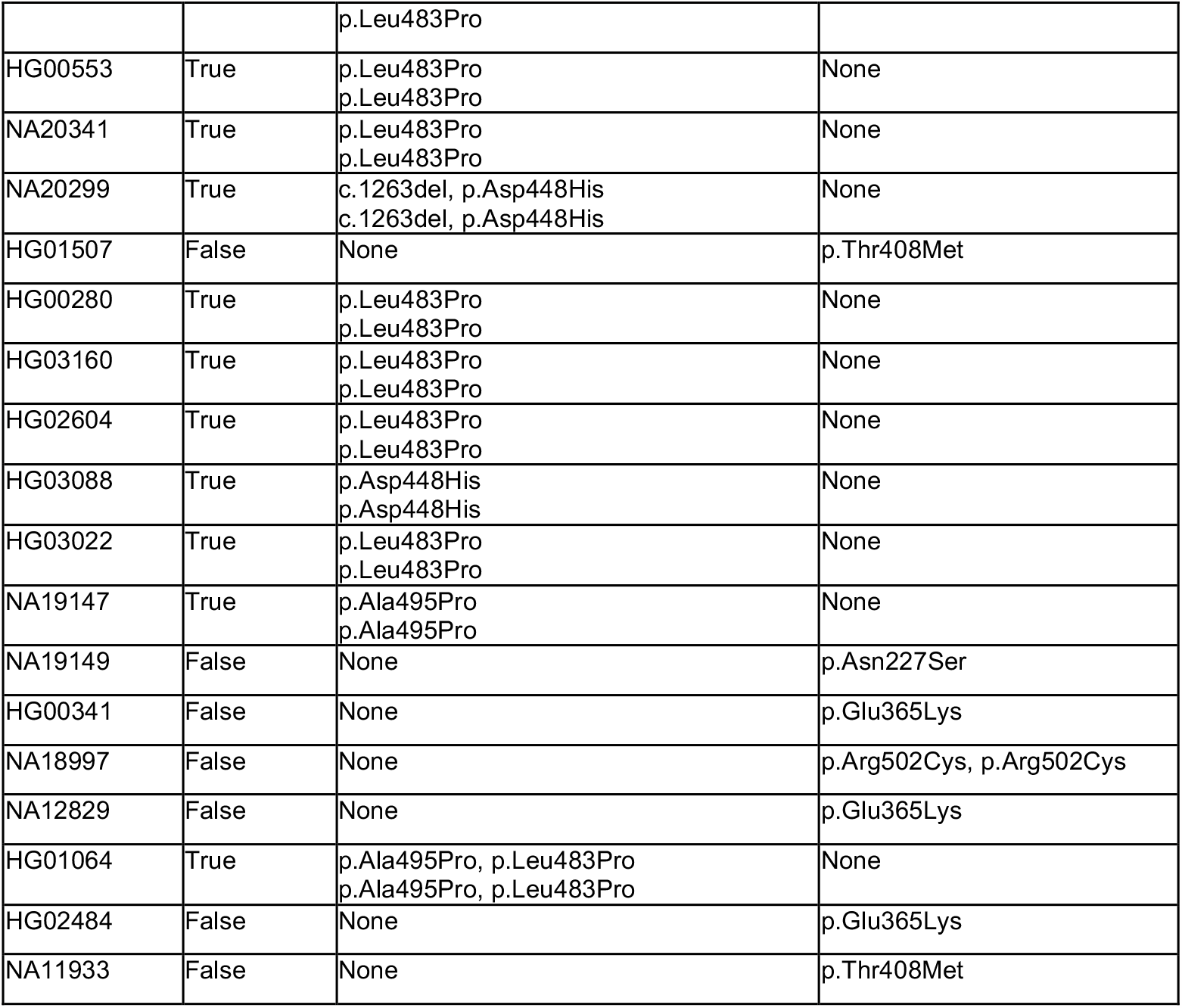
Gauchian *GBA1* variant prediction results for 1000 Genome cases (n=1435). The columns in the Gauchian output for “Carrier”, “CN”, and “*GBA1* Deletion Breakpoint” showed no deviation from “normal” (“False”, “4”, “N/A” respectively) and were omitted from the table.

| Sample | Biallelic | *GBAP1*-like variants in Exons 9-11 | "Unphased" ( *GBA1*-specific)<br>Variants |
| --- | --- | --- | --- |
| HG00327 | True | c.1263del, p.Asp4448His<br>c.1263del, p.Asp4448His | None |
| NA18961 | True | p.Asp448His<br>p.Asp448His | None |
| HG00542 | True | p.Ala495Pro<br>p.Ala495Pro | None |
| HG00136 | True | p.Ala495Pro<br>p.Ala495Pro | None |
| HG00436 | True | p.Asp448His<br>p.Asp448His | None |
| HG01375 | True | p.Ala495Pro<br>p.Ala495Pro | None |
| NA12340 | True | c.1263del, p.Asp448His<br>c.1263del | None |
| NA19197 | True | p.Ala495Pro<br>p.Ala495Pro | None |
| HG01613 | True | p.Asp448His<br>p.Asp448His | None |
| HG02016 | False | None | p.Leu483Arg |
| HG01069 | True | p.Leu483Pro | None |
|  |  | p.Leu483Pro |  |
| HG00553 | True | p.Leu483Pro<br>p.Leu483Pro | None |
| NA20341 | True | p.Leu483Pro<br>p.Leu483Pro | None |
| NA20299 | True | c.1263del, p.Asp448His<br>c.1263del, p.Asp448His | None |
| HG01507 | False | None | p.Thr408Met |
| HG00280 | True | p.Leu483Pro<br>p.Leu483Pro | None |
| HG03160 | True | p.Leu483Pro<br>p.Leu483Pro | None |
| HG02604 | True | p.Leu483Pro<br>p.Leu483Pro | None |
| HG03088 | True | p.Asp448His<br>p.Asp448His | None |
| HG03022 | True | p.Leu483Pro<br>p.Leu483Pro | None |
| NA19147 | True | p.Ala495Pro<br>p.Ala495Pro | None |
| NA19149 | False | None | p.Asn227Ser |
| HG00341 | False | None | p.Glu365Lys |
| NA18997 | False | None | p.Arg502Cys, p.Arg502Cys |
| NA12829 | False | None | p.Glu365Lys |
| HG01064 | True | p.Ala495Pro, p.Leu483Pro<br>p.Ala495Pro, p.Leu483Pro | None |
| HG02484 | False | None | p.Glu365Lys |
| NA11933 | False | None | p.Thr408Met |

Among the 95 cases evaluated, four carried recombinant alleles, two with Rec*NciI*, one with RecTL+55bp and one with a Rec7 allele, which is a rare duplication that includes the *GBA1* gene and duplicated pseudogene [15] (Table 3). Gauchian reported both Rec*NciI* cases correctly (Table 3); however, a detailed analysis of the bam file generated during the alignment of the WGS data for Pat_95 did not identify the known Rec*NciI* mismatches in exon10 (p.Leu483Pro, p.Ala495Pro, and p.Val499Val), but identified 3’UTR mismatches associated with *GBAP1* (Supplemental Figure 2). Visualizing the intron 9 and the 3’-UTR *GBAP1* mismatches indicates a fusion allele (a cross-over between the *GBA1* gene and its pseudogene). In this case a copy of *GBA1P* and the intergenic region is missing. Therefore, only three copies of the gene and pseudogene should be reported, rather than the four copies identified by Gauchian. For the other Rec*NciI* case (Pat_71), only p.Leu484Pro and not the other two mismatches in exon 10 or 3’UTR *GBA1*/*GBAP1* mismatches could be visually identified in the WGS (Supplemental Figure 2). For the second recombinant allele, Rec*TL*+c.1263del (Pat-16, Table 3) a careful analysis of the WGS bam file failed to show the expected exon 9-11 gene/pseudogene mismatches (p.Asp448His, p.Leu483Pro, p.Ala495Pro, and p.Val499Val) (Supplemental Figure 2). Here, Gauchian correctly reported the copy number to be three for this allele, resulting from a cross-over between the gene and pseudogene in intron 8. This generated a fusion-gene which lacked the intergenic region and one copy of the pseudogene (see Supplemental Figure 3). However, Gauchian missed the missense recombinant mutation on the second allele and misreported the heterozygous p.Asn409Ser mutation as homozygous.

Another issue is that Gauchian misreported copy number gains in some cases. This can be seen in the case with the third type of recombinant allele, Rec7, that results in a *GBAP1* duplication (Pat_92, Table 3). This recombination event is rare and is the most difficult recombinant allele to identify [15, 20, 27]. The site of cross-over is between *GBAP1* and *GBA1* or between the contiguous gene Metaxin1 (*MTX1*) and pseudometaxin1 (*MTX1P1*), causing a complete *GBAP1* duplication (Supplemental Figure 2) [20]. Careful evaluation of the WGS from this case demonstrated changes in the read depth of the intergenic area compared to the rest of the genome (Supplemental Figure 2). While other techniques identified one extra copy of the pseudogene, Gauchian indicated three extra copies, reporting a copy number of seven. In two additional cases Gauchian identified increased copy numbers (Pat_42 and Pat_72, Table 3), without signs of a recombinant allele, which requires further investigation.

Another genetic event resulting in variants detected in this cohort is gene conversion. Four patients carried an allele with c1263del (55bp del) in exon 9. While the deletion was genotyped correctly by Gauchian, it identified a gene fusion event (represented as a copy number loss) only in one of the cases (RecTL+55bp in Pat_16).

To further evaluate whether Gauchian may falsely report *GBA1* mutations, the data for 1435 samples from phase 3 of the 1000 Genomes project [28] were extracted and run using Gauchian. Gauchian returned biallelic variants in 20 cases (28 variants total), all called as a *GBAP1*-like variant in exons 9-11, and eight other unphased variants (1.94%) (Table 4, Supplemental Table 5). An analysis of these samples identified no overlap between the predictions and the sample phenotype (WT). Considering both the Gaucher cohort characterized by Sanger sequencing and the 1000 Genomes cohort, Gauchian achieved a sensitivity of 0.89, specificity of 0.98, precision of 0.79, and accuracy of 0.97 (Supplemental Table 6).

## Discussion

Our largest human chromosome, chromosome 1, contains almost 8% of all genetic information. It includes more than 4,220 genes, as well as 991 pseudogenes, of which 840 are processed [29]. Furthermore, several hundred overlapping coding sequences and over 350 disease genes, including mendelian, complex, and neurodegenerative diseases are located on this gene-dense chromosome [29]. Intrachromosomal duplications and deletions of genes and intergenic areas have also been identified, generating gene duplications, recombinant alleles, and structural variations [29, 30]. *GBA1* with its pseudogene, and the neighboring gene *MTX1* with its own pseudogene *MTXP* have a propensity to generate such genomic rearrangements which complicates genotyping of patients with GD (Figure 1). Because they tend to evaluate larger cohorts, PD investigators have sought a means to easily screen their patients to identify those carrying a *GBA1* risk variant. The tool Gauchian was designed to provide this service, and it can accurately identify many *GBA1* variants. However, it still does not overcome the challenges posed by the presence of complex recombination in this locus. Here we show that even when Gauchian accurately identified recombinant alleles, it could not demonstrate the underlying mechanism of recombination. For example, Rec*NciI* may result from gene fusion or gene conversion, and each may have a different effect on phenotype. We also found that WGS alone could not accurately describe many of the recombinant alleles. Currently a combination of approaches is needed to accurately detect and define recombinant alleles. Perhaps the use of long-read WGS will ultimately be helpful.

Another concern is that even with missense mutations, Gauchian identified some heterozygote missense mutations as homozygous changes, which can have important implications for patients and their families. Of further relevance is an internal discrepancy in the performance of Gauchian when using difference human reference sequences. While existing benchmarking approaches comparing the nature of small variants between the reference sequences hg19 and hg38 have favored the newer hg38 [26], Gauchian performs better when running on the older reference version for most common genotypes. This adds further complexity when applying Gauchian as a diagnostic tool. Additionally, due to the limited number of variants in its database, the software may incorrectly report a carrier or even a patient with GD as normal, which again can have clinical repercussions and can distort PD data analysis. We especially recommend caution when using Gauchian to evaluate newborns or children suspected of having GD. Future studies should specifically compare the results of Sanger sequencing, copy number analyses, and WGS with Gauchian predictions in larger cohorts, including more individuals with known complex alleles.

Our attempt to use Gauchian to evaluate WGS data from the 1000 Genomes Project is also concerning. While we do not have Sanger sequence data on the 1435 individuals evaluated by Gauchian, it is highly unlikely that twenty would have homozygous mutations leading to previously undiagnosed GD, especially as the majority of variants identified were *GBAP1* mismatches located in exons 9-11. Moreover, most of these predicted variants are associated with type 3 GD, which usually presents in childhood and is not likely to be found in a control adult cohort.

Furthermore, the tool is unable to find *de novo* mutations, since it can only detect variants in its database of 121 known *GBA1* variants, and three known recombination events. Since to date over 500 *GBA1* SNV variants and at least three additional recombinant alleles have been described [12], this is a further limitation of the tool. With the wider use of both whole *GBA1* and next generation sequencing, more variants are continually being identified.

In conclusion, while Gauchian may have value in a research setting, we highly recommend that any results used for clinical purposes be validated with Sanger sequencing or at least using another means to evaluate the WGS. Furthermore, Gauchian should not be used as a standalone blackbox test in larger diagnostic variant calling pipelines like Dragen4 for the purpose of high-throughput genotyping, or as the basis for patient counseling.

## Methods

High molecular weight DNA was extracted from 95 patients (90 with GD and five known carriers of *GBA1* variants) seen under an NIH Institutional Review Board-approved natural history study with informed consent. Each sample underwent both Sanger sequencing and WGS, performed at the NIH Intramural Sequencing Center.

*GBA1-*related genotypes for each patient were established using Sanger sequencing with primers and protocols introduced in Stone et al. [31]. WGS sequencing libraries were generated from 1μg genomic DNA using a TruSeq® DNA PCR-Free HT Sample Preparation Kit (Illumina, #FC-121-3001), with median insert sizes of approximately 400 bp. Libraries were tagged with barcodes to allow pooling and were sequenced on the NovaSeq 6000 (Illumina, RRID:SCR_016387) to obtain at least 300 million 151-base read pairs per individual library. The bioinformatics pipeline followed guidelines for the Genome Analysis Toolkit (GATK, RRID:SCR_001876) [32]. Reads were aligned to the human b37_decoy reference sequence (UCSC assembly hg19, NCBI build 37) using BWA (RRID:SCR_010910, http://bio-bwa.sourceforge.net/ accessed on 22 September 2022) [33]. Gauchian (v.1.0.2, https://github.com/Illumina/Gauchian accessed on 18 March 2022) [25] was run using the b37 reference genome.

To evaluate the performance of Gauchian against the assembled cohort of patients with GD, we used Sanger calls for a specific known mutation as positive observations across the afflicted alleles and the absence of the same mutation in an allele as negative observations. For example, if a patient was observed to have a genotype p.N409S/p.N409S, the p.N409S variant would be assigned 2 positive observations and 0 negative ones, while a p.L483P variant would be assigned 0 positive observations and 2 negative ones for this patient. After mapping of all the variants observed across the cohort, it is then possible to categorize each Gauchian prediction (or absence of prediction) into 1) a true positive call, where Gauchian detected the correct Sanger variant, 2) a true negative call, where Gauchian correctly omitted a variant, 3) a false positive call, where Gauchian predicted a variant in the patient, but Sanger and visual inspection of the WGS contradicted the finding, and 4) a false negative call, where Gauchian did not report a variant that was expected based on Sanger sequencing. With the total set of true positives and negatives, as well as false positives and negatives it is then possible to characterize the overall performance using metrics like sensitivity, specificity, accuracy and precision.

Additionally, to further evaluate the frequency of possible false positives, WGS data from phase 3 of the 1000 Genomes project [28] was evaluated by Gauchian. To streamline performance, samples were retrieved as sets of 100, and were directly processed in Gauchian utilizing the computational resources of the NIH HPC Biowulf cluster (http://hpc.nih.gov). The results of the 25 individual Gauchian runs were merged into a single output table and filtered for calls containing relevant *GBA1* variants, indicated through explicitly predicted *GBAP1*-like variants in exons 9-11, or for *GBA1*-specific variants.

## Statistics and Reproducibility

No statistical analyses were used to explore the data due to the nature of the provided analysis. The data necessary to reproduce the analysis is provided and shall be used together with Gauchian version 1.0.2.

## Data Availability

The data is being prepared for the submission to dbGaP and an accession number will be provided as soon as it is available.

Acknowledgments

This work was supported by the intramural research programs of the National Human Genome Research Institute and the National Institutes of Health. This research was also funded in part by Aligning Science Across Parkinson’s [ASAP-000458] through the Michael J. Fox Foundation for Parkinson’s Research (MJFF). For the purpose of open access, the author has applied a CC-BY public copyright license to all Author Accepted Manuscripts arising from this submission.

The authors would like to thank Andrew Hogan, Andrea D’Souza, Geena Woo and the NIH Intramural Sequencing Center (NISC) for the preparation and sequencing of the samples, and Emory Ryan and Dr. Grisel Lopez for collecting the clinical samples.

## Author Contributions

NT coordinated the project, organized the sequencing, analyzed the results, and drafted the manuscript, JL conducted the data analysis and drafted the manuscript. EH contributed to the interpretation of the data. ES supervised the execution of the project, data interpretation and the preparation of the manuscript. All authors have read and approved the manuscript.

## Supplemental Tables

Supplemental Table 1. The complete set of 95 whole genome sequencing samples was processed using Gauchian and the results are presented with the original headers provided by the tool. Sanger-established genotype and gender are provided for the NIH cohort and a final assessment comparing the Gauchian call against the Sanger genotype is presented.

Supplemental Table 2. Breakdown of Gauchian performance based on the individual alleles. For each allele, Sanger sequencing was used to determine the total number of occurrences of a genotype across all 190 alleles (4 alleles have two variants) as positive identifications and the number of alleles without the genotype as negative identifications. The same approach was used to group the Gauchian predictions into positive and negative calls. Based on these allele specific groupings it is then possible to identify true and false positives as well as true and false negatives. Finally, the sum of incorrectly identified alleles is provided for each genotype as well as the sums for the different metrics for the complete cohort, which corresponds to the entries in a statistical classification specific confusion matrix.

Supplemental Table 3. Derivations for an allele specific confusion matrix. Each of the metrics presented is based on ratios of total, true or false positive and negative counts or other confusion matrix derivations.

Supplemental Table 4. Comparison of Gauchian using b37 and hg38 as reference genomes against the established Sanger calls.

Supplemental Table 5. The complete set of 95 whole genome sequencing samples from the NIH cohort as well as 1435 whole genome sequencing samples extracted from the 1000 Genomes project was processed using Gauchian. The results are presented here using the original headers provided by the tool. Each Gauchian genotype was compared against the calls established for the NIH cohort and the 1000 Genomes Project. The comparison of the two genotypes was then broken down into true positive identification and true negative identifications, and each cohort-specific genotype was characterized as a positive call (biallelic GBA1 mutation), negative call (biallelic wildtype), or a combination of the two.

Supplemental Table 6. Derivations for confusion matrix considering the NIH Cohort, 1000 Genomes Cohort as well as the two cohorts combined. Each of the metrics presented is based on ratios of total, true or false positive and negative counts or other confusion matrix derivations.

## Supplemental Figures

Supplemental Figure 1. Visualization of whole genome sequencing results for affected exons in patients containing a mismatch between Sanger/WGS validation and the Gauchian prediction.

Supplemental Figure 2. Visualization of whole genome sequencing results in patients exhibiting recombinant events.

## References

1. Davidson, B.A., et al., Exploring genetic modifiers of Gaucher disease: The next horizon. Human Mutation, 2018. 39(12): p. 1739–1751.

2. Smith, L.J., et al., Genetic variations in GBA1 and LRRK2 genes: Biochemical and clinical consequences in Parkinson disease. Front Neurol, 2022. 13: p. 971252.

3. Neumann, J., et al., Glucocerebrosidase mutations in clinical and pathologically proven Parkinson’s disease. Brain, 2009. 132(Pt 7): p. 1783–94.

4. Chang, D., et al., A meta-analysis of genome-wide association studies identifies 17 new Parkinson’s disease risk loci. Nat Genet, 2017. 49(10): p. 1511–1516.

5. Sidransky, E., et al., Multicenter Analysis of Glucocerebrosidase Mutations in Parkinson’s Disease. New England Journal of Medicine, 2009. 361(17): p. 1651–1661.

6. Horowitz, M., et al., The human glucocerebrosidase gene and pseudogene: structure and evolution. Genomics, 1989. 4(1): p. 87–96.

7. Purandare, S.M. and P.I. Patel, Recombination hot spots and human disease. Genome Res, 1997. 7(8): p. 773–86.

8. Martinez-Arias, R., et al., Sequence Variability of a Human Pseudogene. Genome research, 2001. 11: p. 1071–85.

9. Parks, M.M., C.E. Lawrence, and B.J. Raphael, Detecting non-allelic homologous recombination from high-throughput sequencing data. Genome Biology, 2015. 16(1): p. 72.

10. Tayebi, N., et al., 55-Base pair deletion in certain patients with Gaucher disease complicates screening for common Gaucher alleles. American Journal of Medical Genetics, 1996. 66(3): p. 316–319.

11. Cormand, B., et al., A new gene-pseudogene fusion allele due to a recombination in intron 2 of the glucocerebrosidase gene causes Gaucher disease. Blood Cells Mol Dis, 2000. 26(5): p. 409–16.

12. Hruska, K.S., et al., Gaucher disease: mutation and polymorphism spectrum in the glucocerebrosidase gene (GBA). Hum Mutat, 2008. 29(5): p. 567–83.

13. Koprivica, V., et al., Analysis and classification of 304 mutant alleles in patients with type 1 and type 3 Gaucher disease. Am J Hum Genet, 2000. 66(6): p. 1777–86.

14. Wafaei, J.R. and F.Y.M. Choy, Glucocerebrosidase recombinant allele: Molecular evolution of the glucocerebrosidase gene and pseudogene in primates. Blood Cells, Molecules, and Diseases, 2005. 35(2): p. 277–285.

15. Tayebi, N., et al., Reciprocal and nonreciprocal recombination at the glucocerebrosidase gene region: implications for complexity in Gaucher disease. Am J Hum Genet, 2003. 72(3): p. 519–34.

16. Jeong, S.-Y., et al., Identification of a novel recombinant mutation in Korean patients with Gaucher disease using a long-range PCR approach. Journal of Human Genetics, 2011. 56(6): p. 469–471.

17. Díaz-Font, A., et al., Gene rearrangements in the glucocerebrosidase-metaxin region giving rise to disease-causing mutations and polymorphisms. Analysis of 25 Rec NciI alleles in Gaucher disease patients. Hum Genet, 2003. 112(4): p. 426–9.

18. Zampieri, S., et al., GBA Analysis in Next-Generation Era: Pitfalls, Challenges, and Possible Solutions. The Journal of Molecular Diagnostics, 2017. 19(5): p. 733–741.

19. Zeng, Q., et al., A customized scaffolds approach for the detection and phasing of complex variants by next-generation sequencing. Sci Rep, 2020. 10(1): p. 15060.

20. Woo, E.G., N. Tayebi, and E. Sidransky, Next-Generation Sequencing Analysis of GBA1: The Challenge of Detecting Complex Recombinant Alleles. Front Genet, 2021. 12: p. 684067.

21. Tayebi, N., et al., Gaucher disease and parkinsonism: a phenotypic and genotypic characterization. Mol Genet Metab, 2001. 73(4): p. 313–21.

22. Velayati, A., et al., Identification of recombinant alleles using quantitative real-time PCR implications for Gaucher disease. J Mol Diagn, 2011. 13(4): p. 401–5.

23. Zhou, Y., et al., Mutational spectrum and clinical features of GBA1 variants in a Chinese cohort with Parkinson’s disease. NPJ Parkinsons Dis, 2023. 9(1): p. 129.

24. Gabbert, C., et al., GBA in Parkinson’s disease: variant detection and pathogenicity scoring matters. medRxiv, 2023: p. 2023.02.03.23285410.

25. Toffoli, M., et al., Comprehensive short and long read sequencing analysis for the Gaucher and Parkinson’s disease-associated GBA gene. Communications Biology, 2022. 5(1): p. 670.

26. Pan, B., et al., Similarities and differences between variants called with human reference genome HG19 or HG38. BMC Bioinformatics, 2019. 20(2): p. 101.

27. Tayebi, N., et al., Genotypic heterogeneity and phenotypic variation among patients with type 2 Gaucher’s disease. Pediatr Res, 1998. 43(5): p. 571–8.

28. Genomes Project, C., et al., A global reference for human genetic variation. Nature, 2015. 526(7571): p. 68–74.

29. Gregory, S.G., et al., The DNA sequence and biological annotation of human chromosome 1. Nature, 2006. 441(7091): p. 315–21.

30. Sønderby, I.E., et al., 1q21.1 distal copy number variants are associated with cerebral and cognitive alterations in humans. Transl Psychiatry, 2021. 11(1): p. 182.

31. Stone, D.L., et al., Glucocerebrosidase gene mutations in patients with type 2 Gaucher disease. Hum Mutat, 2000. 15(2): p. 181–8.

32. DePristo, M.A., et al., A framework for variation discovery and genotyping using nextgeneration DNA sequencing data. Nat Genet, 2011. 43(5): p. 491–8.

33. Li, H. and R. Durbin, Fast and accurate short read alignment with Burrows-Wheeler transform. Bioinformatics, 2009. 25(14): p. 1754–60.

