## Supplemental Figures for "Is Gauchian genotyping of *GBA1* variants reliable?"

**Supplemental Figure 1**

The complete set of 95 whole genome sequencing samples was processed using Gauchian and the results are presented with the original headers provided by the tool. Sanger-established genotype and gender are provided for the NIH cohort and a final assessment comparing the Gauchian call against the Sanger genotype is presented.

A – Pat_03

Sanger Annotation: N370S/N370S

Gauchian Call: N370S/WT

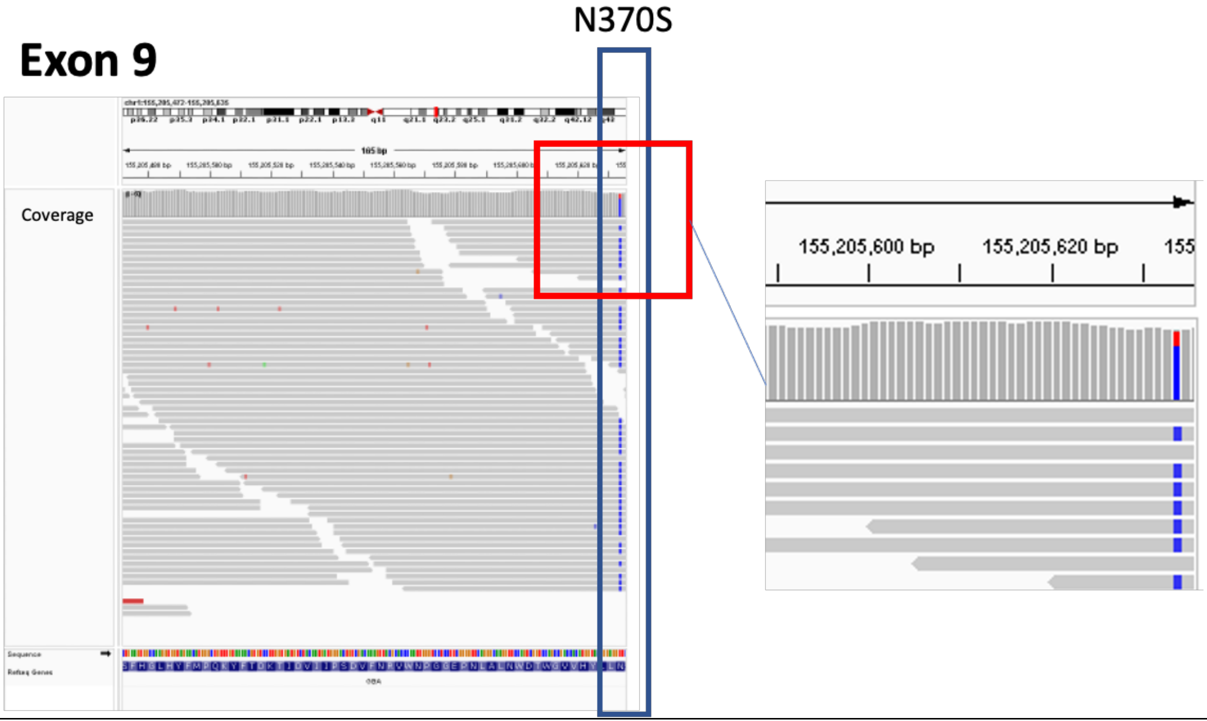


B – Pat_08

Sanger Annotation: N370S/Q350X

Gauchian Call: N370S/WT

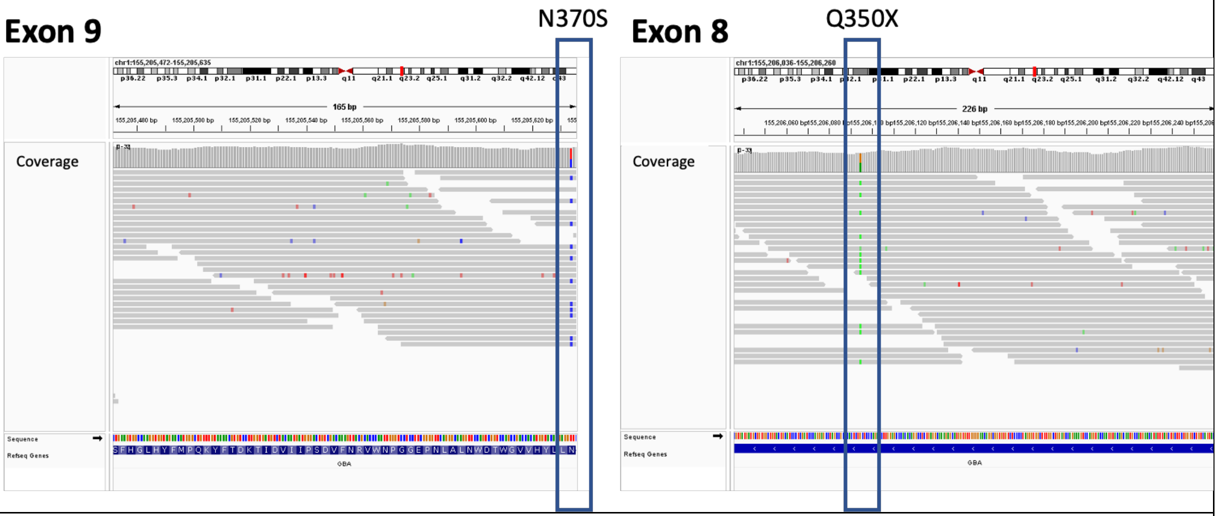


C – Pat_26

Sanger Annotation: N370S/R463C
Gauchian Call: N370S/WT

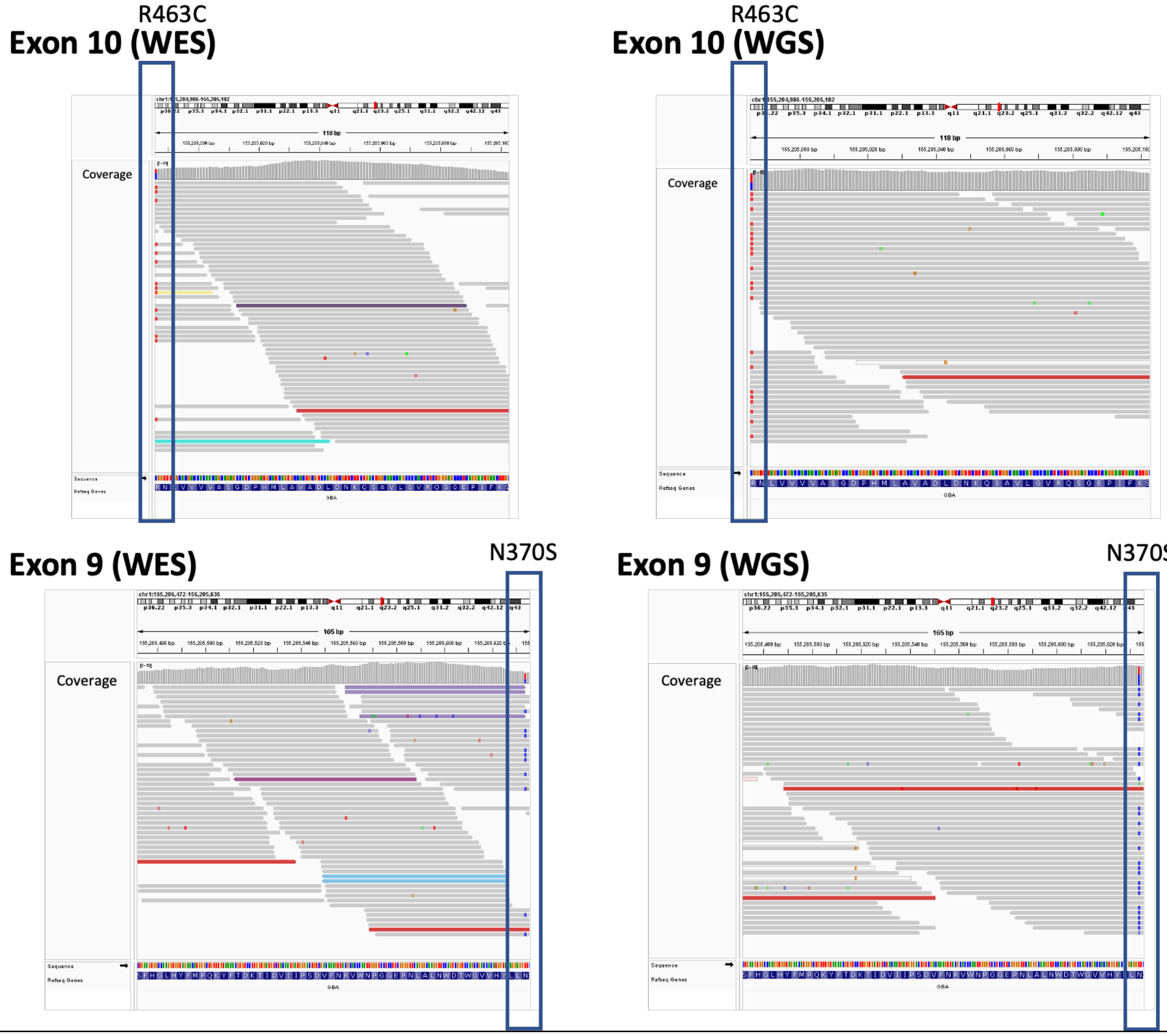


D – Pat_28
Sanger Annotation: R496H/C342Y
Gauchian Call: R496H/WT

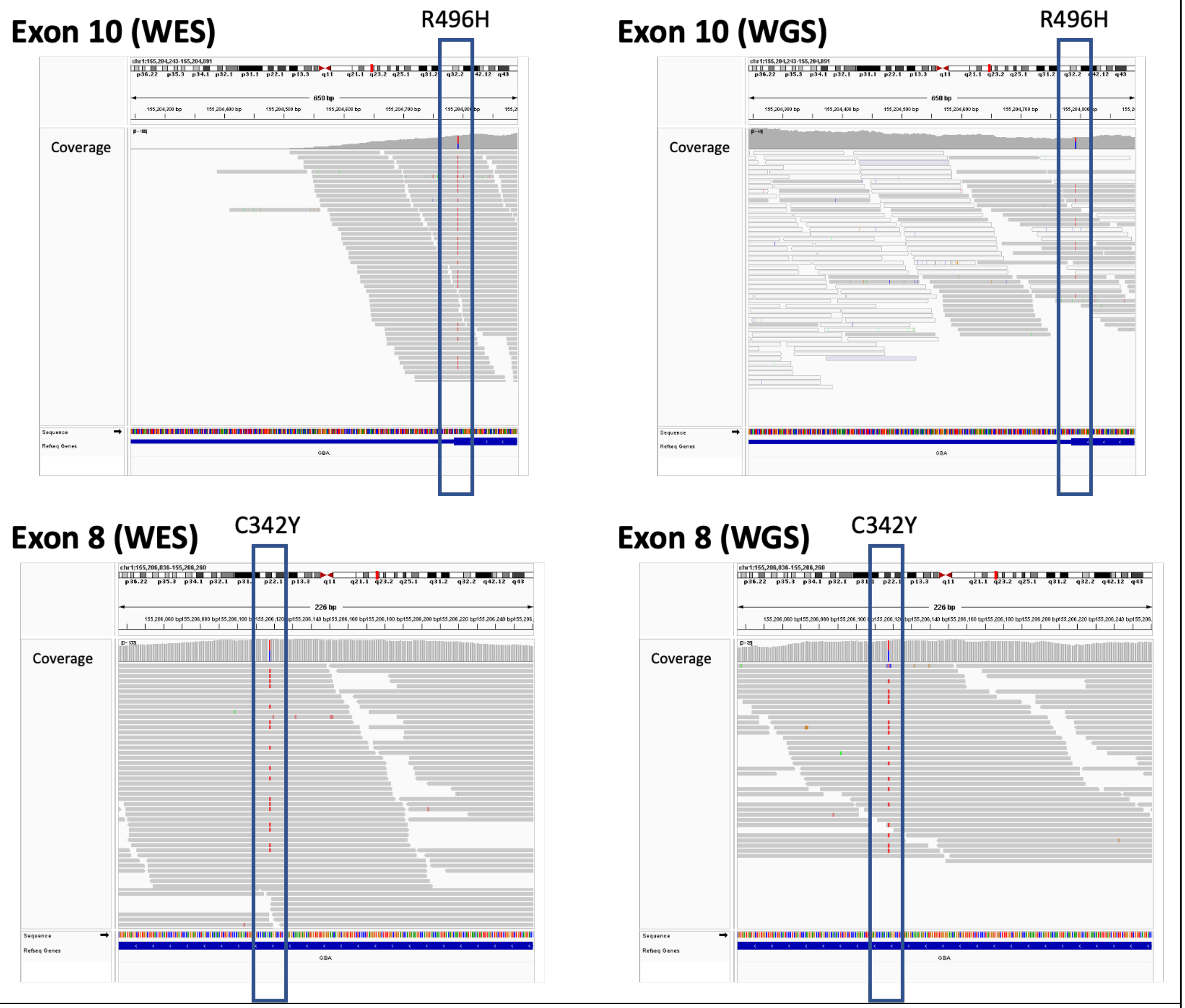


E – Pat_47

Sanger Annotation: N370S/L444P

Gauchian Call: N370S/WT

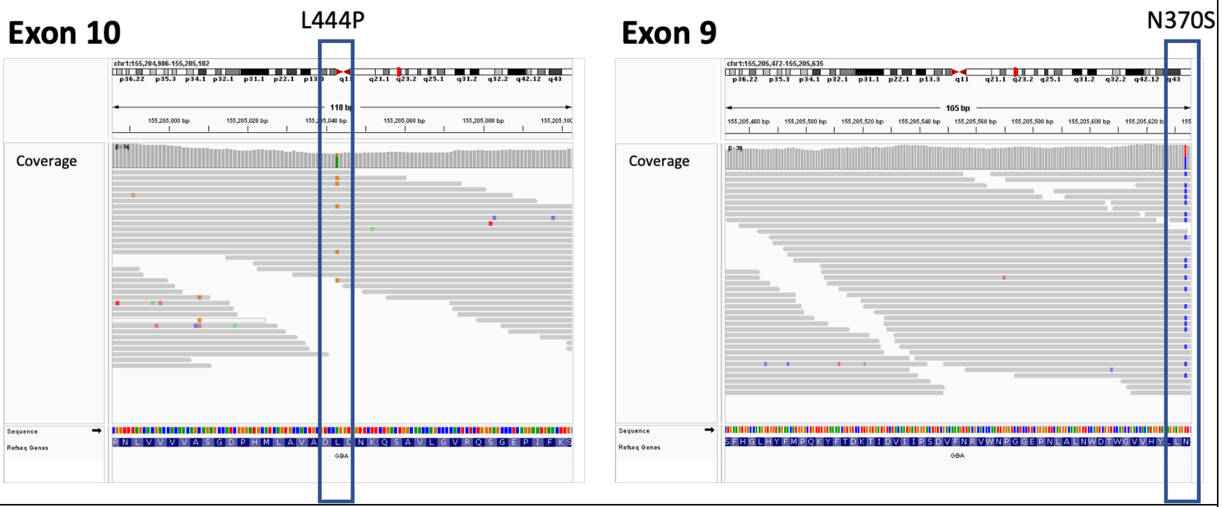


F – Pat_58

Sanger Annotation: N370S/R257*/c203delC
Gauchian Call: N370S/R257*

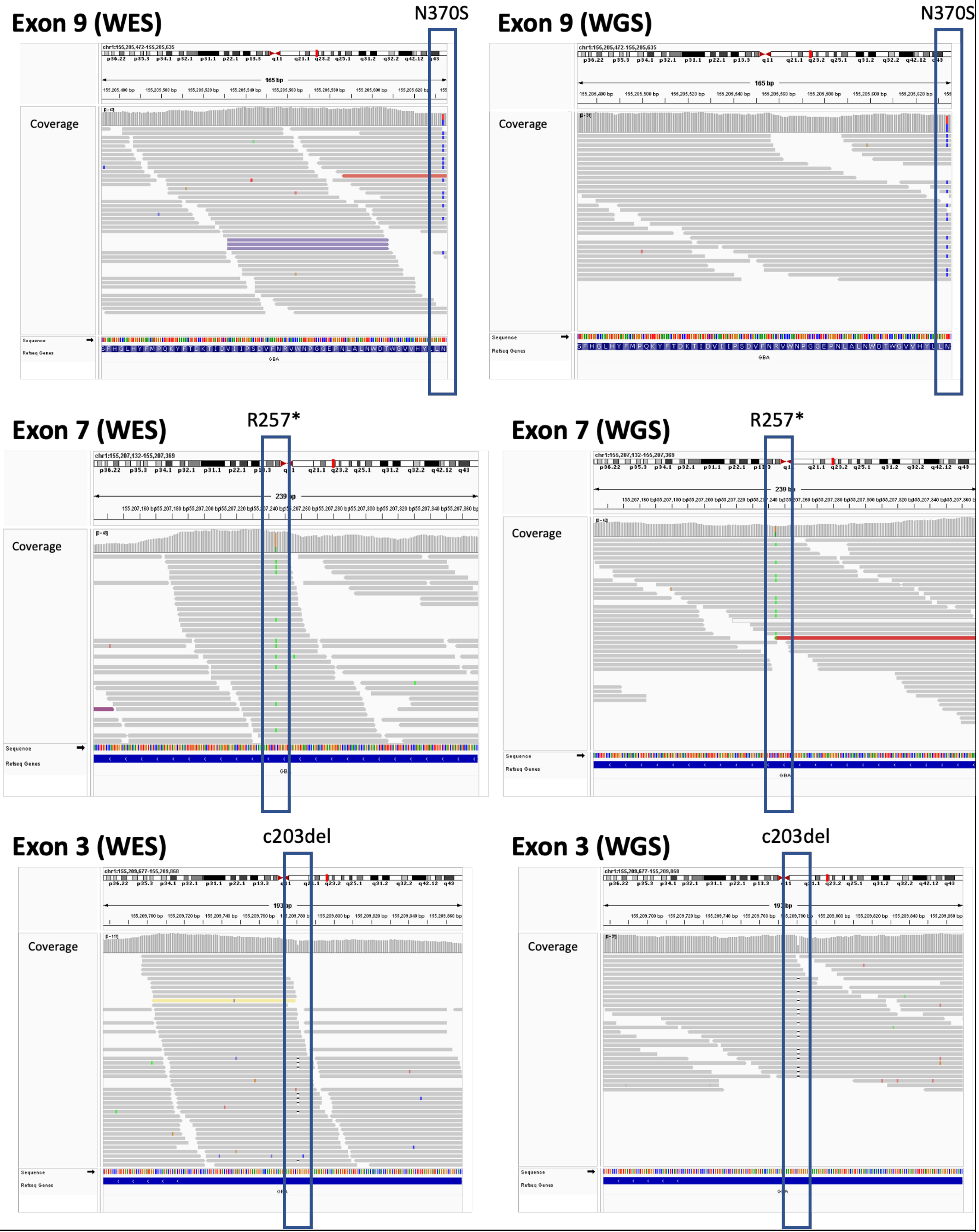


G – Pat_75
Sanger Annotation: R463C/R120W

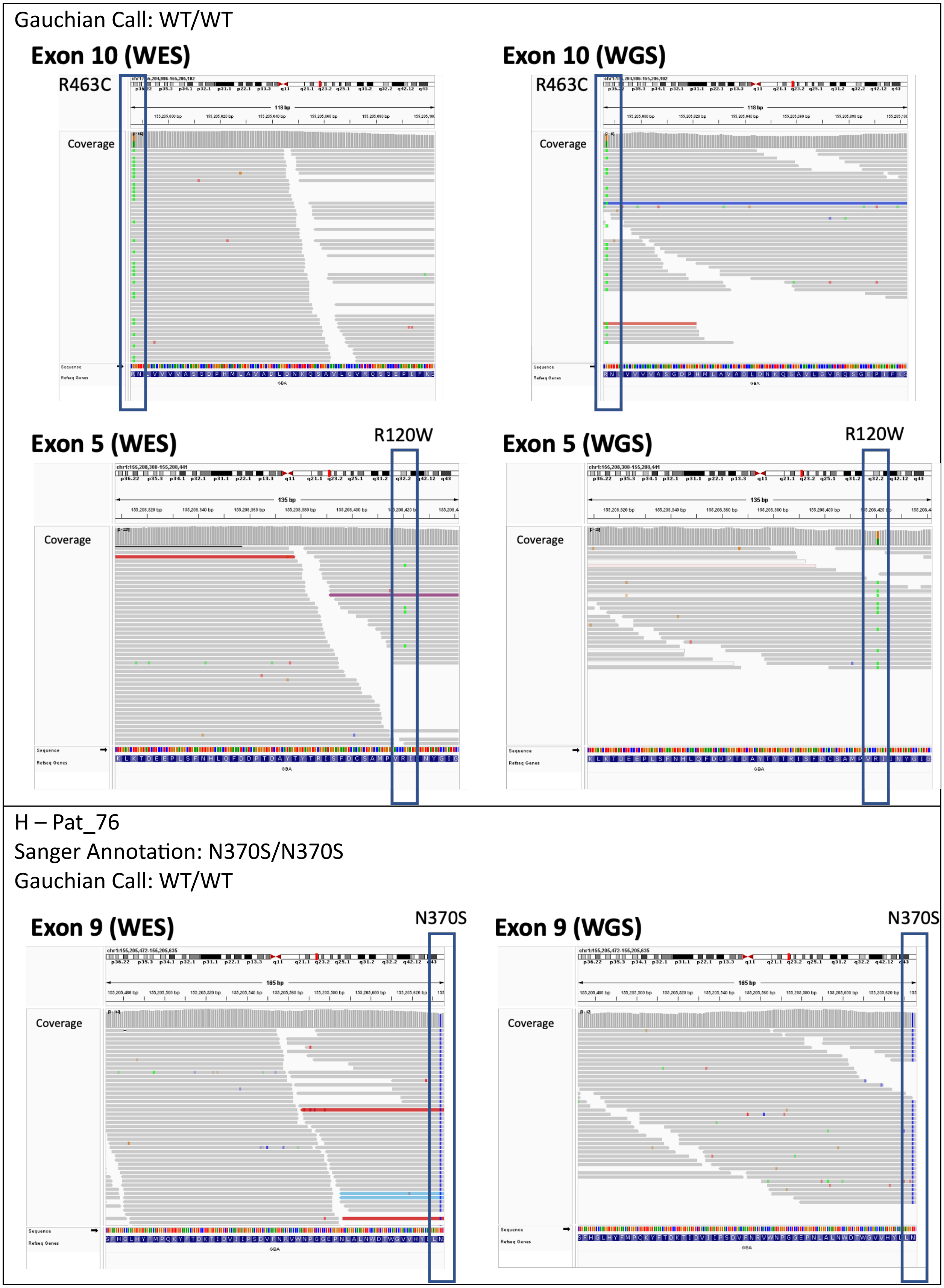
Gauchian Call: WT/WT
Exon 10 (WES)
R463C
Coverage
Exon 10 (WGS)
R463C
Coverage
Exon 5 (WES)
R120W
Coverage
Exon 5 (WGS)
R120W
Coverage
H – Pat_76
Sanger Annotation: N370S/N370S
Gauchian Call: WT/WT
Exon 9 (WES)
N370S
Coverage
Exon 9 (WGS)
N370S
Coverage

I – Pat_79

Sanger Annotation: L444P/R463C

Gauchian Call: WT/WT

**Exon 10 (WGS)**

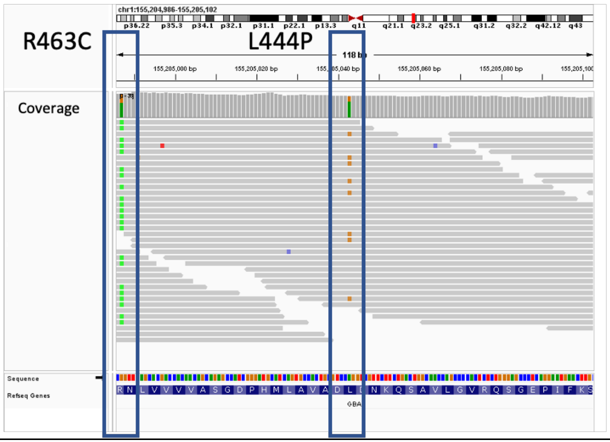


J – Pat_92

Sanger Annotation: D409H/L444P + Rec7

Gauchian Call: D409H/D409H,L444P

**Exon 9**

D409H

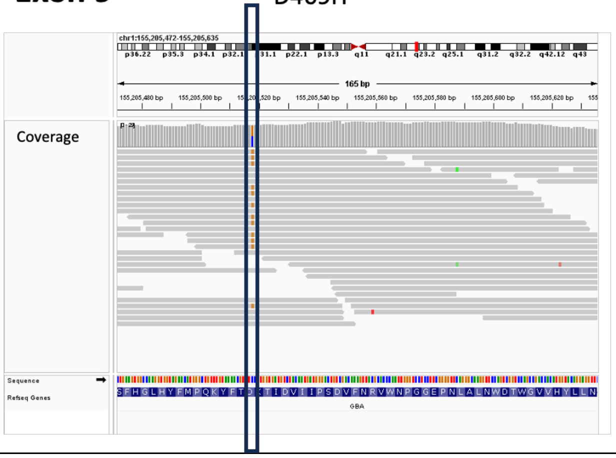


**Exon 10**

L444P

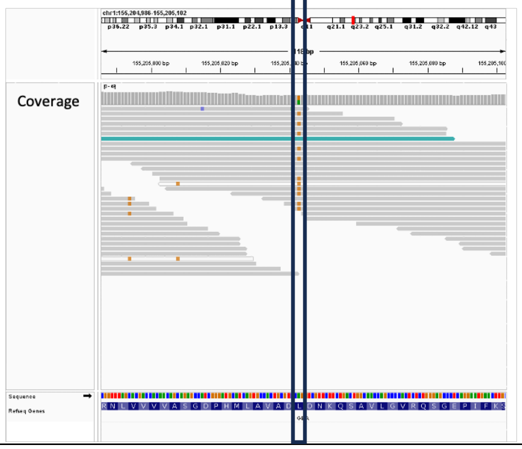

**Supplemental Figure 2**

Visualization of whole genome sequencing results in patients exhibiting recombinant events.

A – Pat_95 (RecNciI/p.Asn409Ser)

Exon 9 shows a heterozygous mutation for p.Asn409Ser, while exon 10 shows that the variants associated with RecNciI are not explicitly marked and exon 11 shows that the GBA1/GBAP1 SN mismatch is detectable.

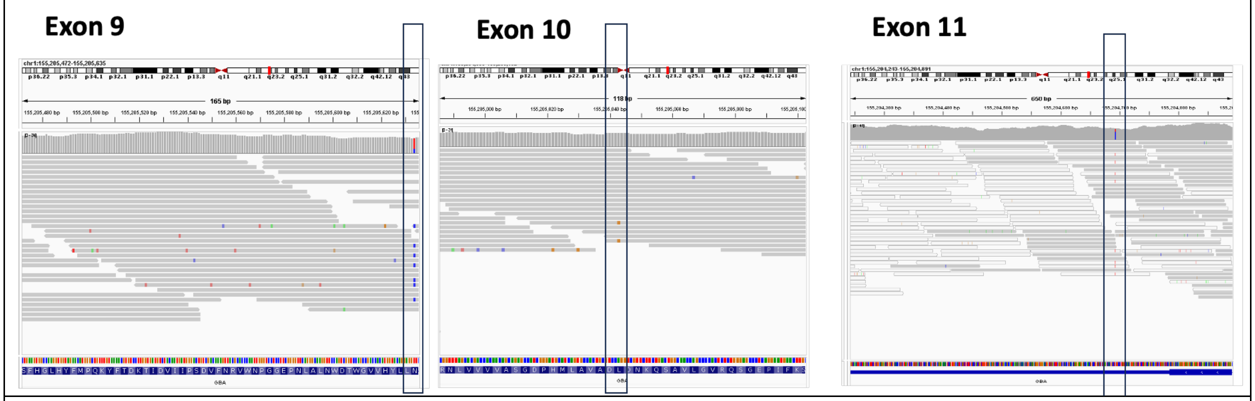


B – Pat_71 (RecNciI/WT)

Exon 10 shows only p.Leu483Pro detectable for the RecNciI variants and the expected mismatches in the 3'-UTR and Intron 9 are undetectable.

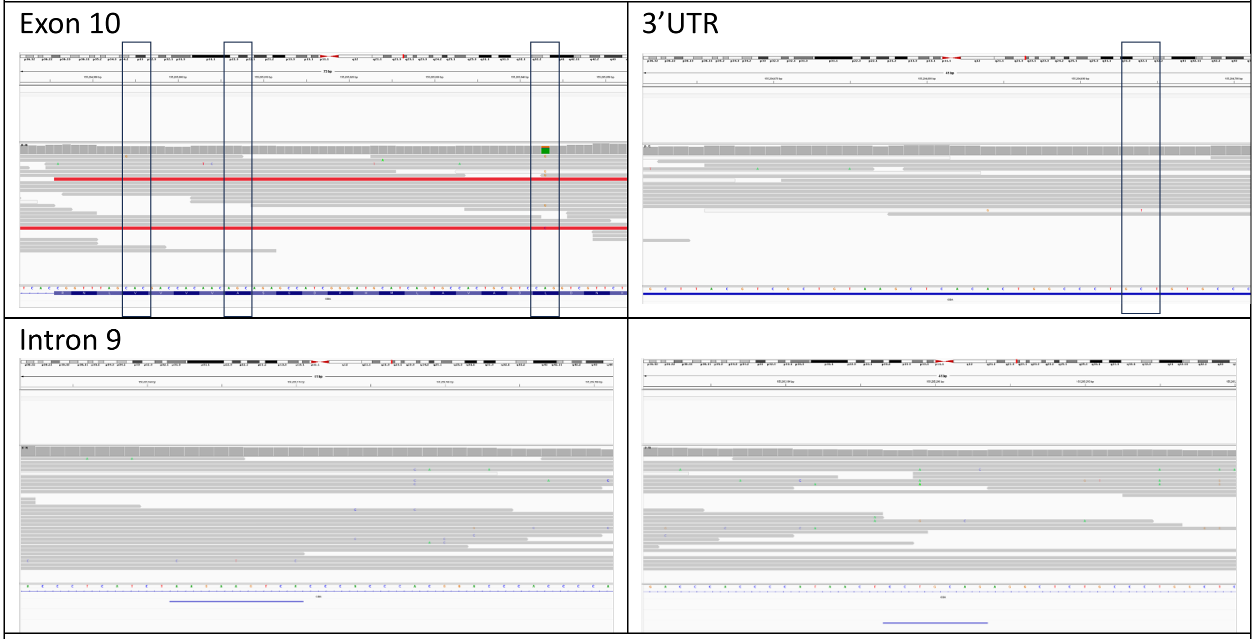


C – Pat_16 (p.Asn409Ser,RecTL+55bpdel)

Exon 9 shows detectable p.Asn409Ser, 55bp deletion but no p.Asp448His. Exon 10 shows that none of the three expected variants are detectable, with the expected mismatches in the 3'UTR also not detectable.

Exon 9

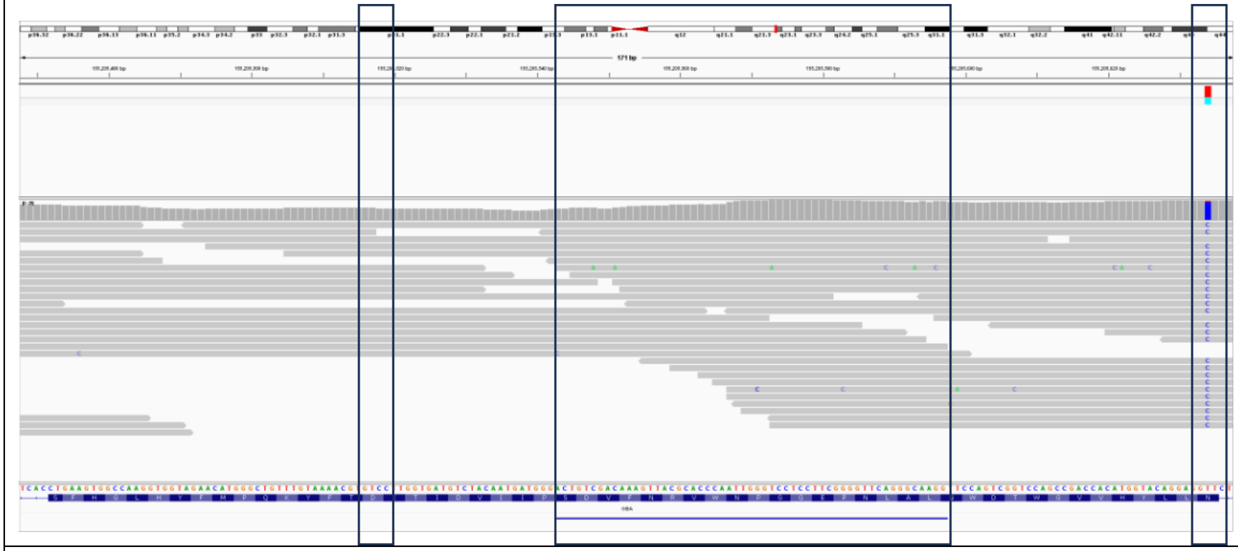

Exon 10

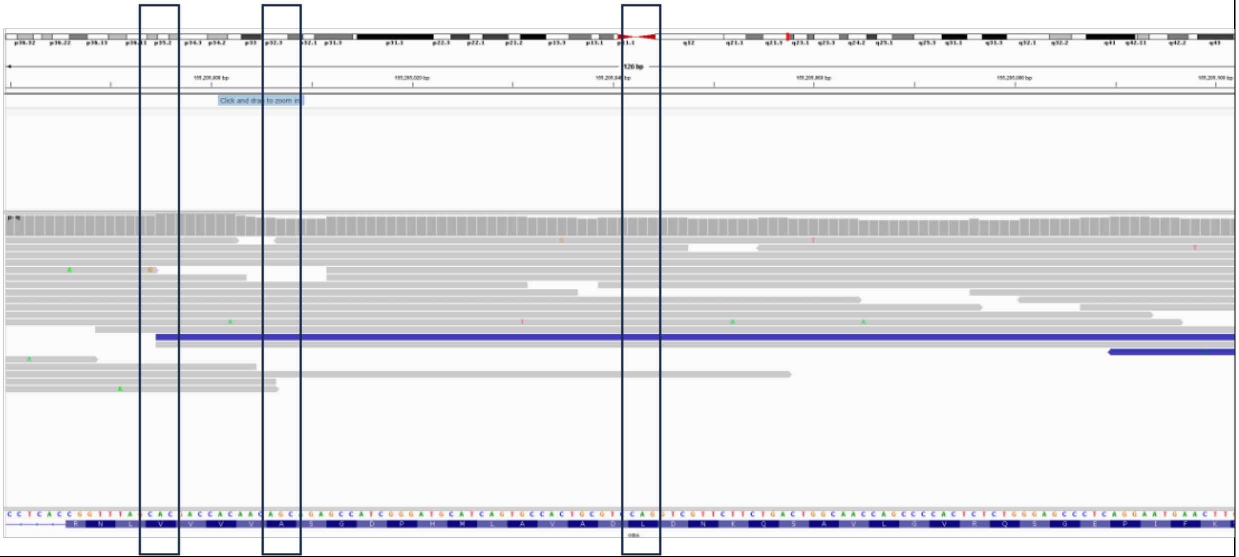

3'UTR

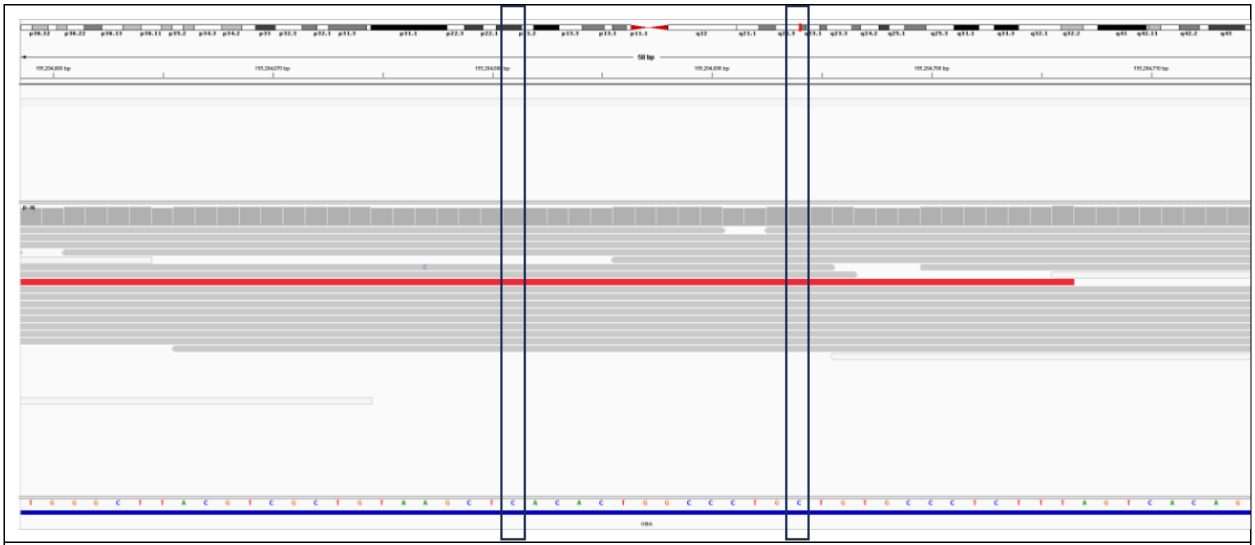

D – Pat_92 (p.Asp448His/p.Leu483Pro, Rec7)
For this *GBA1P* duplication, Gauchian predicted three extra copies (CN=7). The box highlights the possible duplicated area and both missense mutations were detectable.

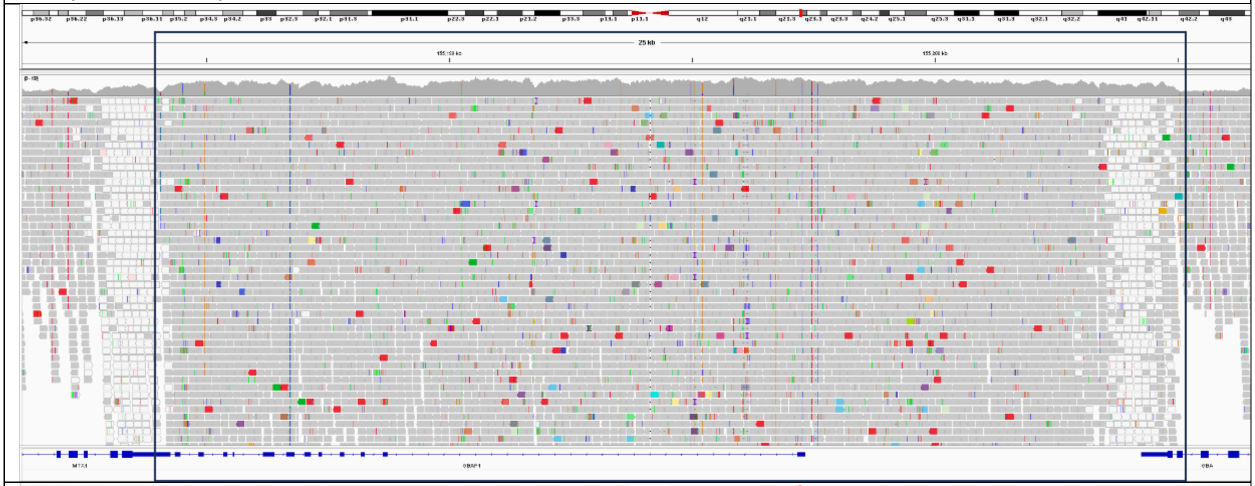

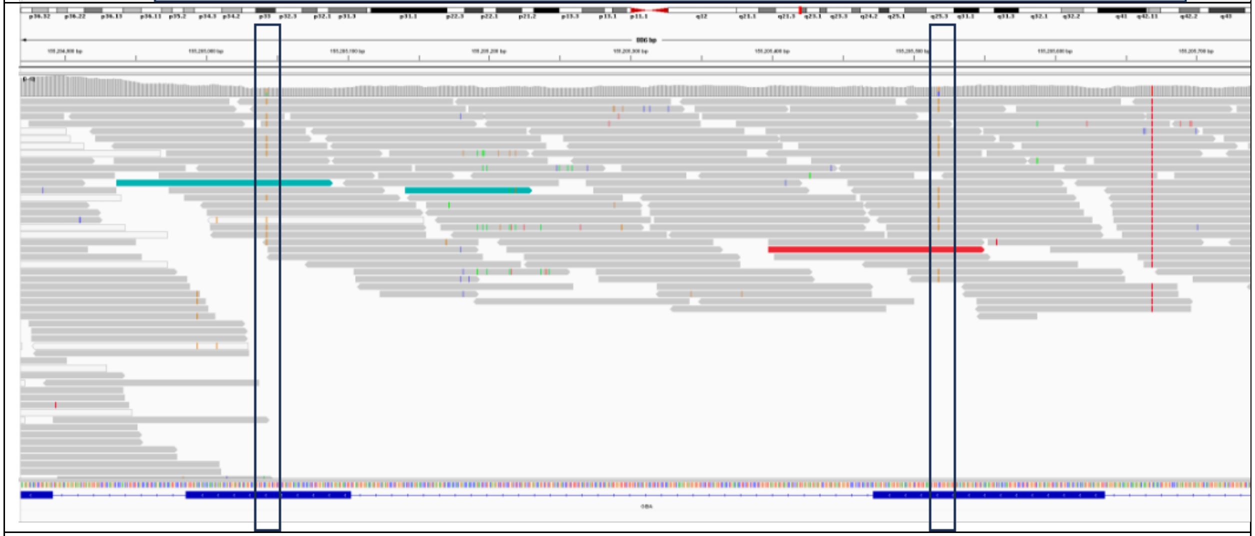

E – Pat_42 (p.Val391Leu/p.Arg398Ter)

For this patient Gauchian predicted two extra copies and a duplication can be detected for *GBAP1*.

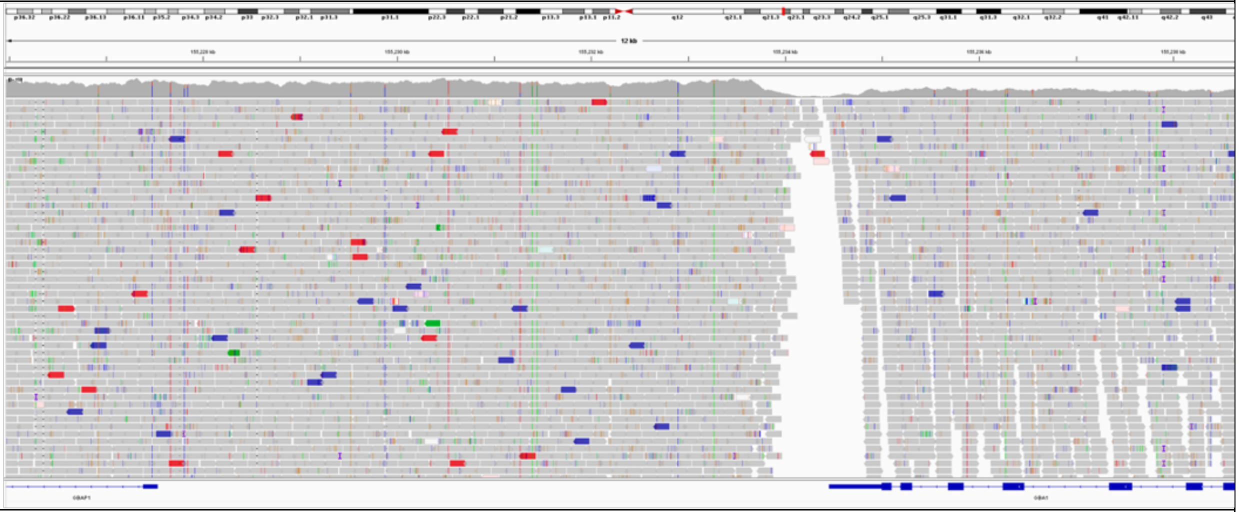

F – Pat_72 (p.Gly241Arg/WT)

For this patient Gauchian predicted one extra copy, which can also be detected for *GBAP1*.

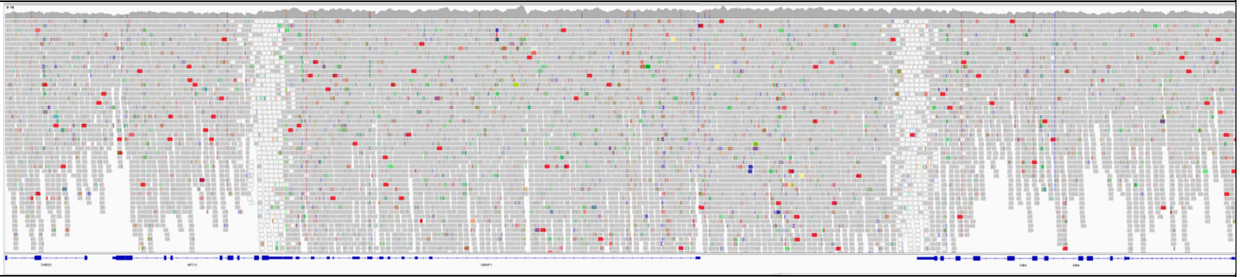

G – Pat_15 (55bpdel/p.Asn409Ser)

The 55bp deletion and the heterozygous p.Asn409Ser mutation in exon 9 are detectable.

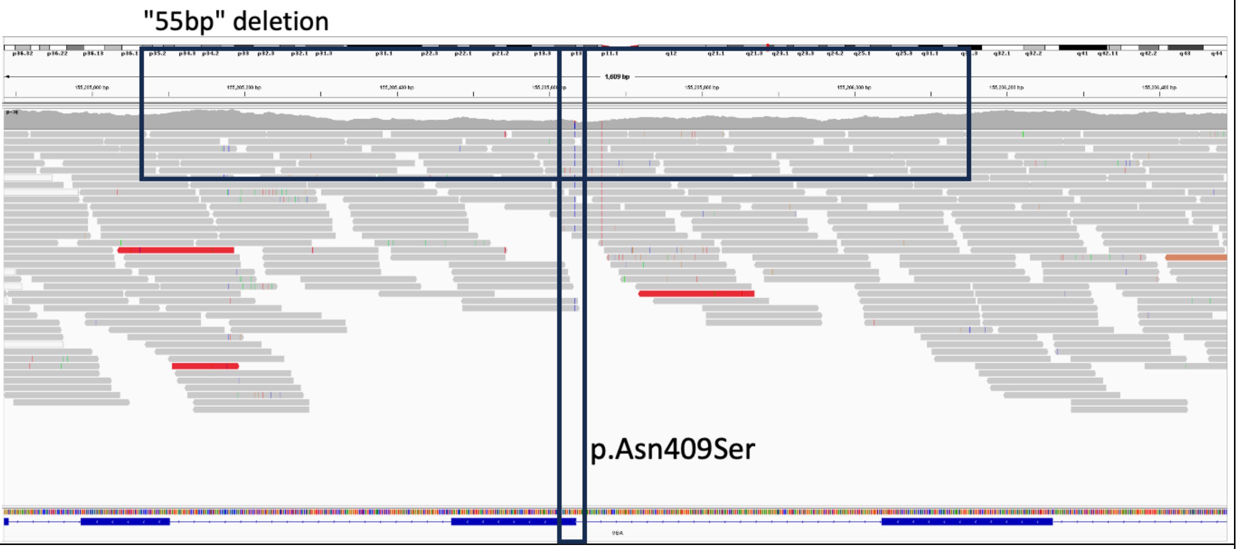


H – Pat_39 (55bpdel/p.Asn409Ser)

The 55bp deletion and the heterozygous p.Asn409Ser mutation in exon 9 are detectable.

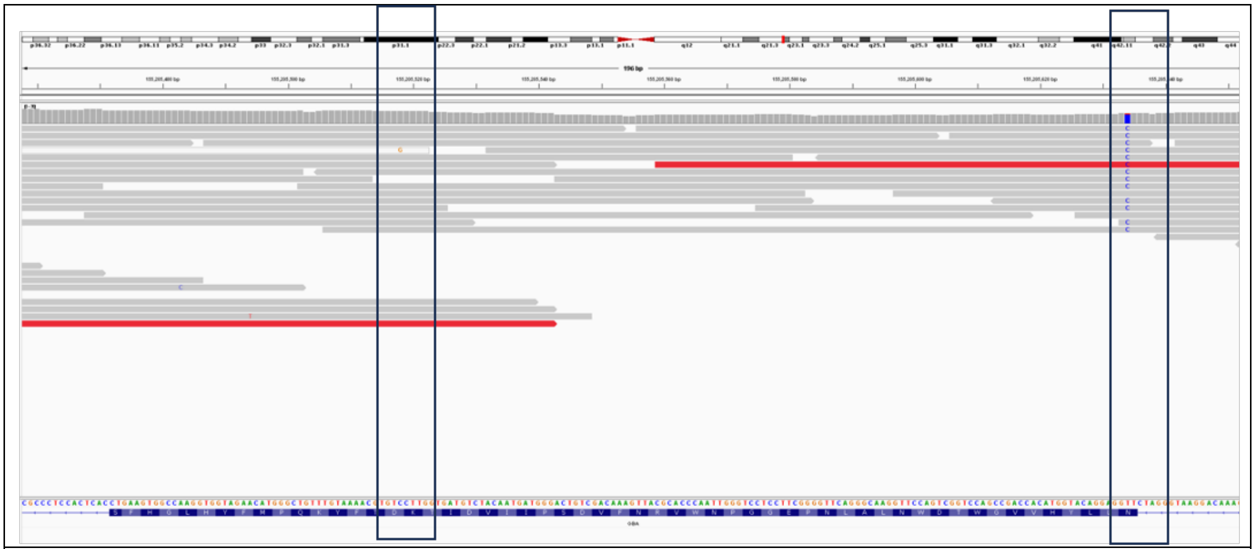

I – Pat_69 (55bpdel/WT)
The 55bp deletion in exon 9 is detectable.

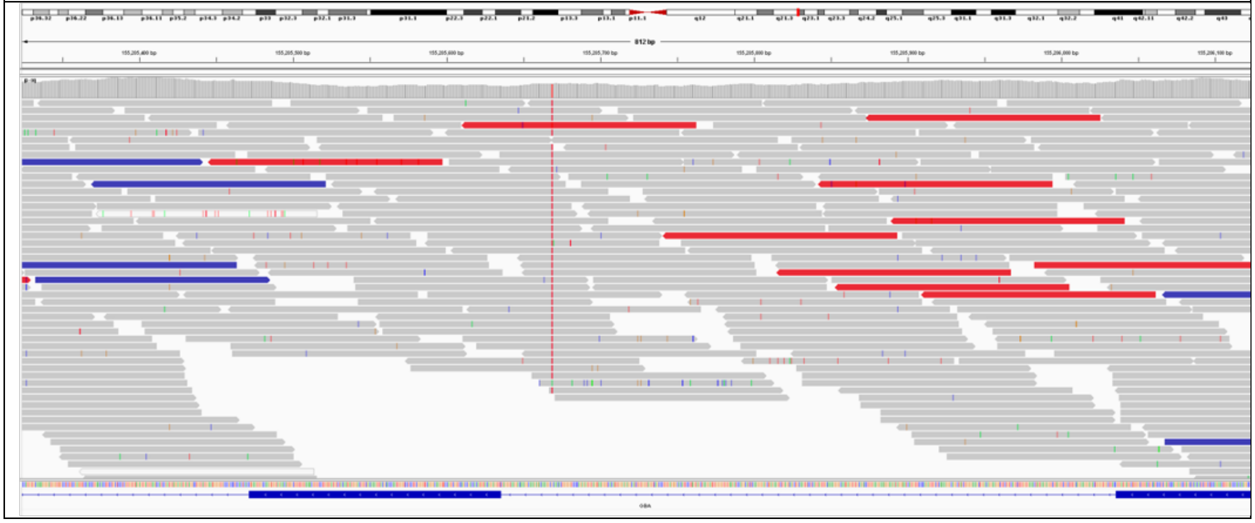
